# Systematic Review and Meta-analysis of Eculizumab, Inebilizumab, Tocilizumab, and Satralizumab for Neuromyelitis Optica

**DOI:** 10.1101/2021.07.22.21261005

**Authors:** Rajan Chamlagain, Sangam Shah, Suman Gaire, Anuj Krishna Paudel, Krishna Dahal, Bipin kandel, Roman Dhital, Basanta Sharma Paudel, Sandesh Dhakal, Madan Basnet

## Abstract

Neuromyelitis optica is rare, autoimmune-mediated inflammation and demyelination of the central nervous system with a prevalence of 1-2 persons per 100,000 populations. We aim to generate a head-to-head comparison of these drugs with appropriate evidence to guide future trials and treatment guidelines in a patient with recurrent attacks of NMO. We searched the databases like PubMed, MEDLINE, Cochrane Central Register of Controlled Trials (CENTRAL) and Embase for studies published prior to April 2021 using the keywords. Over all 929 patients from 11 different publications were included in the study. Five studies were included for quantitative synthesis. Pooling of studies showed significant mean reduction of ARR in the monoclonal antibody group (-0.26 [-0.35, -0.17], P <0.00001, I^2^=0%) and the mean difference in EDSS score from baseline in monoclonal antibodies was - 0.23(95% CI [-0.43, -0.03], P=0.02, I^2^=0%). There was no significant difference in frequency of total reported adverse events between monoclonal antibody and the comparator arm (RR: 1.01 [0.95, 1.07], P=0.74, I^2^=14%). Our findings, particularly seen from the context of a few RCTs, support the pursuit of larger, multi-center RCTs that evaluate the effectiveness of each of the currently available monoclonal antibodies and better describe their adverse risk profile.

## Introduction

Neuromyelitis optica is a rare, autoimmune-mediated inflammation and demyelination of the central nervous system with a prevalence of 1-2 persons per 100,000 populations.^1,2^ It predominately affects women in the ratio of 9:1.^3^ NMO is a monophasic or relapsing-remitting disorder predominantly characterized by optic neuritis and transverse myelitis.^4^ Optic nerve involvement presents with blindness or vision loss, motor impairment, sensory disorder, urination/defecation function disturbance, and vomiting due to the attack of the spinal cord, brain stem involvement resulting in intractable nausea are some main features of a patient with NMOSD.^5–8^ Transverse myelitis presents with longitudinally extensive spinal cord lesions; a tendency to spare the brain, but when the brain is affected, the presence of magnetic resonance imaging (MRI) lesions is atypical for MS; and frequent association with seropositivity for NMO IgG (IgG antibody to aqua-porin-4). The key marker in NMOSD is an aquaporin-4 antibody (AQP4-Ab) and accounts for 80% of NMOSD cases.^9^ Myelin oligodendrocyte glycoprotein antibody (MOG-Ab) discovered recently is another biomarker and is found in 4-11% NMOSD patients but does not co-exist with AQP4-Ab seropositivity.^10^

The pathophysiologic process of NMO is complex involving B cell-mediated production of pathological autoantibody, immunoglobulin G (IgG) which mainly targets the astrocyte water channel aquaporin-4 (AQP4).^11,12^ AQP4-IgG binds to astrocytic AQP4 and triggers classical complement cascade activation, promotes granulocytic and lymphocytic infiltration which will then combine to damage neural tissues.^13^ IL-6 plays a pivotal role in driving the disease activity by stimulating AQP4-IgG secretion and plasmablast survival, disrupting blood-brain barrier integrity and production of proinflammatory T-lymphocyte and their differentiation and activation.^14^ IL-6 levels are elevated in the serum and cerebrospinal fluid of patients with NMO.^14,15^ Hence, the main goals in treatment of NMO include acute symptomatic therapy and long-term prevention of relapses. For acute management, corticosteroids and/or plasmapheresis are used while drugs like rituximab, mycophenolate, and azathioprine are recommended for maintenance therapy.^16–18^ These drugs might be effective in preventing relapses but the prolonged or even lifelong immunosuppression often leads to inevitable adverse effects. Hence, newer therapies (eculizumab, inebilizumab, tocilizumab, and satralizumab) are proposed to prevent future attacks in patients of neuromyelitis optica.^2,19^ These therapies possess different targets within the immune pathogenic process and hence, can alter the outcome in patient of neuromyelitis optica.^2^

Eculizumab inhibits the classical complement system and formation of membrane attack complex thus, inhibiting neuronal injury.^20^ Inebilizumab is another humanized, binds to the B-cell surface antigen CD19 that identifies and depletes a wider range of lymphocytes exclusively from the B-cell lineage.^21^ In contrast, satralizumab and tocilizumab are humanized monoclonal antibody that binds to IL-6 receptors and inhibits the IL-6 signalling pathways involved in inflammation.^22,23^ However, these drugs are still not compared based on decreased relapse risk, Annualized Relapse Rate (ARR) ratio, improve Expanded Disability Status Scale (EDSS) score, serious adverse events, and mortality. Therefore, in this study, we aim to generate a head-to-head comparison of these drugs with appropriate evidence to guide future trials and treatment guidelines in a patient with recurrent attacks of NMO.

### Literature Review

#### Methods

This meta-analysis confirms to standard guidelines and is written in accordance with Preferred Reporting Items for Systematic Reviews and Meta-Analyses guidelines (PRISMA) statement^24^. (CRD42021255886)

### Study selection

We performed comprehensive and systematic literature search for all studies about the use of eculizumab, inebilizumab, satralizumab, and tocilizumab to treat NMOSD patients. We searched the databases like PubMed, MEDLINE, Cochrane Central Register of Controlled Trials (CENTRAL) and Embase for studies published prior to April 2021 using the keywords ‘neuromyelitis optic spectrum disorders’ or ‘NMOSD’ or ‘aquaporin 4 antibody’, or ‘devic’s disease, or ‘monoclonal antibody’ and the drug names ‘tocilizumab’, ‘eculizumab’, ‘inebilizumab’, and ‘satralizumab’. We also searched the reference lists of all included studies and any associated review articles to identify any relevant studies that were missed in the initial search. The detail study of study selection is shown by flowchart in Fig. 1.

**Figure 1:**
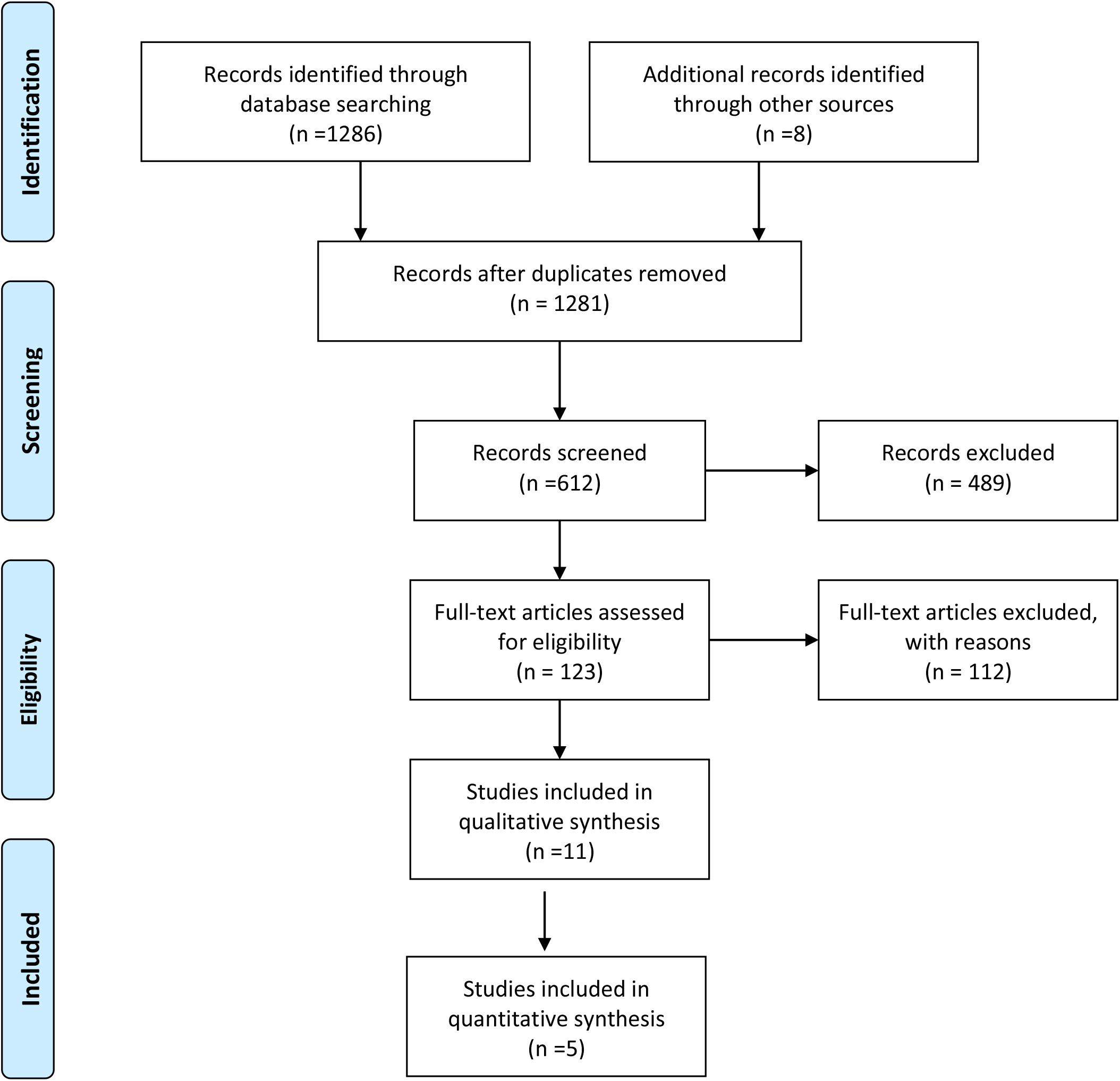
PRISMA guidelines for article identification and selection

Two reviewers (SS and RC) independently screened the retrieved articles and obtained the full texts of all the articles that met the predefined selection criteria. Any discrepancies between reviews were solved by discussion with the third author (BSP, KD, SG, AKP). We read full studies after identifying all the articles and screening the abstracts. Ultimately, twelve studies met the inclusion criteria and were included in the review (Supplemetal table 1).

### Inclusion and Exclusion criteria

We included the studies that fulfilled the following criteria: (1) The study population composed of NMOSD patients with any of its types (2) Treatment with tocilizumab, or eculizumab, or inebilizumab, or satralizumab; (3) The articles that were available in English language and was of human. The exclusion criteria were as follows: (1) case reports and studies that included less than ten patients; (2) studies that were published in other languages (3) data presented only in posters, abstracts, or presentations.

For quantitative analysis only the randomised control trials that included tocilizumab, eculizumab, inebilizumab, or satralizumab in their intervention arm and placebo or immunosuppressive agents in their control arm were included.

### Data extraction

Two authors (SS and RC) independently extracted the data from the included studies and any discrepancies were solved by discussion with other authors (BSP and KD). The following information was extracted from the studies: the author’s name and year of publication, study design, country where the study was done, treatment drugs used, AQP4-IgG serostatus, history of relapse, history of rituximab, dosage regimen, sample size, follow up duration, and efficacy measures.

### Main variables

We assessed three primary efficacy outcome measures: (1) Annualized Relapse Rate (ARR) ratio, (2) mean EDSS score and (3) pain and fatigue severity scale. Safety outcomes included the proportion of deaths and adverse effects.

### Quality assessment

We evaluated following items in the assessment: (1) clarity of the study objectives; (2) whether the study period (start date and end date) was stated clearly? (3) whether the description of the patient selection criteria was clear ? (4) study was multinational or multicentric; (5) Treatment with any of the drug (Eculizumab, or Inebilizumab, or Satralizumab, or Tocilizumab) and its dose; (6) whether the baseline equivalence groups were clearly considered ; (7) the definition of the primary outcome (annualised relapse rate, change in EDSS score and Pain and fatigue severity scale) prior to the study; (8) if the follow-up period was long enough (months); (9) whether a clear hazard ratio (HR) with 95% confdence intervals (95% CI) was stated and (10) the limitations of each study. We did not use quality assessment as exclusion criteria. According to the quality items used in each study (score range 0–10), the papers were assessed.

### Synthesis of results

Revman 5.4 was used for statistical analysis of the extracted data. Mean difference was used to evaluate the outcome in case of continuous data. Random effects model was used for the synthesis of result. The dichotomous outcomes were evaluated as risk ratio by using random effects model. The outcomes were reported along with 95 % confidence interval and a P value (<0.05 being significant). Forest plots were used for graphical representation of outcome. I^2^ test was used to measure the statistical heterogeneity. The test was interpreted as: 0% to 40%: might not be important; 30% to 60%: may represent moderate heterogeneity; 50% to 90%: may represent substantial heterogeneity; 75% to 100%: considerable heterogeneity as per Cochrane handbook of systematic reviews^25^. Subgroup and sensitivity analysis were not performed.

## Results

### Literature search

A total of 1294 studies were found in the initial electronic searches from PubMed, Cochrane Library, Embase, and clinicaltrials.gov. 123 studies were included in the full text screening which after reviewing the included papers carefully, 112 articles were excluded leaving 11 paper for qualitative synthesis (Tocilizumab n=5, Satralizumab n=2, Ecuilizumab n=3, and Inebulizumab n=1). The details of the study selection are as shown in Figure 1. The quality of the 11 included papers was fair with an average score of 8.72 and a median score of 8 (range 7-10) (Supplemental table 1).

### Literature details

Over all 929 patients from 11 different publications were included in the study. Three of the studies assessed patients from Germany, each two from USA and Japan, four studies were from multiple countries and one from China. Nine studies were prospective in design, of which 6 were randomized control trials while three studies were retrospective in design. Five studies included patients with positive AQP4 serostatus while other included patients with both positive and negative AQP4 serostatus. All studies included patients with positive history of relapse while 1 study hadn’t taken it as inclusion criteria. Five studies included the patients who had taken previous rituximab therapy. The detail of the characteristics of the included study is summarized in table 1.

### Qualitative analysis

The outcomes of the included studies have been listed in the following table 2.

### Quantitative analysis

We included 5 RCTs in our quantitative analysis.

### Outcomes

#### 1. Relapse

The risk of relapse was evaluated by difference in ARR and number of on-trial relapse in monoclonal antibody arm versus the comparator arm. Only three studies reported difference in ARR whereas all included RCTs reported number of on-trial relapses.

Pooling of three studies showed significant mean reduction of ARR in the monoclonal antibody group (−0.26 [-0.35, -0.17], P <0.00001, I^2^=0%).Similarly, pooling of data from all included RCTs showed a lower relapse risk in the monoclonal antibody group as compared to control (RR: 0.32 [0.19, 0.55], P<0.0001). However, the heterogeneity in the on trial risk was significant I^2^=69%. (Figure 2)

**Fig 2:**
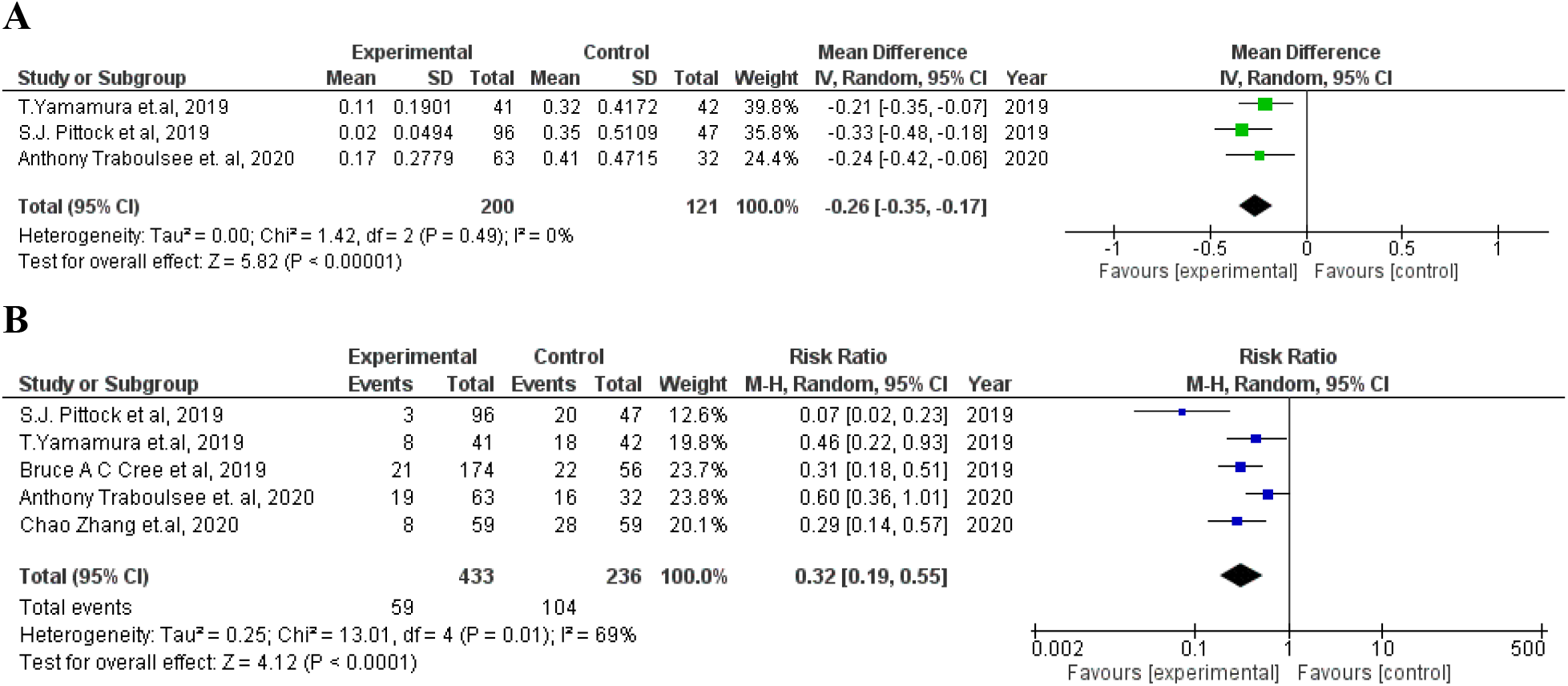
A: Change in ARR B. On trial relapse

#### 2. Change in EDSS score

All studies contained information on EDSS score, however there was incomplete reporting or different form of reporting. So, data regarding change in EDSS score from baseline was pooled from three studies. The mean difference in EDSS score from baseline in monoclonal antibodies was -0.23(95% CI [-0.43, -0.03], P=0.02, I^2^=0%) (Figure 3)

**Figure 3:**
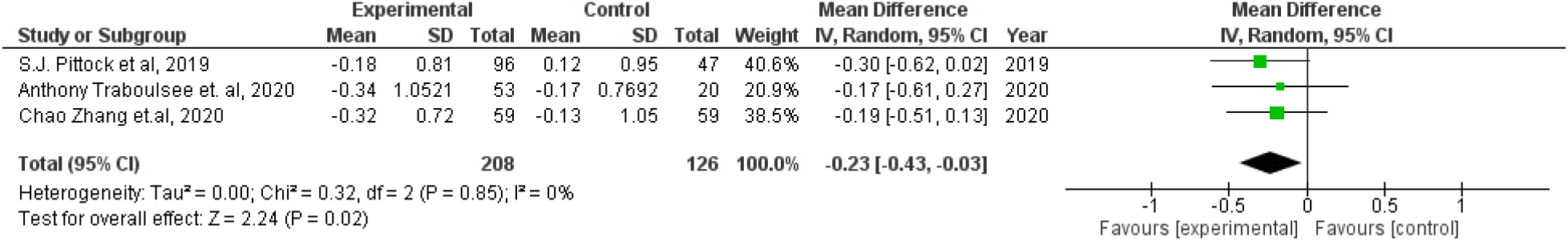
A: Change in EDSS score

#### 2. Pain score and fatigue severity score

Change in VAS pain score was reported in 2 scores. Pooling of change in VAS pain score from 2 RCTs showed the mean difference of pain score in monoclonal antibodies to be 4.06 (95% CI: [2.22, 5.90], P<0.0001, I^2^=0%). (Figure 4A) Pooling of change in FACIT-F score from 2 studies did not show significant mean difference in FACIT-F fatigue score (−1.02 [-6.02, 3.97], P=0.69). However, there was significant heterogeneity in the finding I^2^=78%). (Figure 4B)

**Figure 4:**
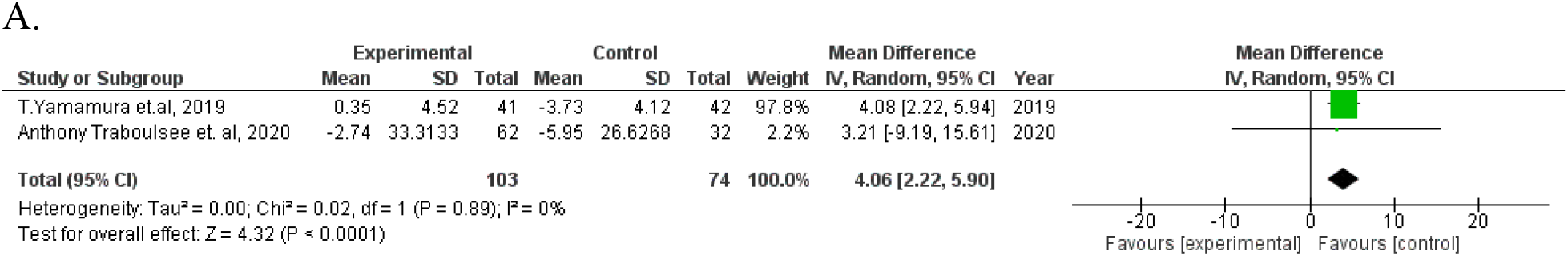
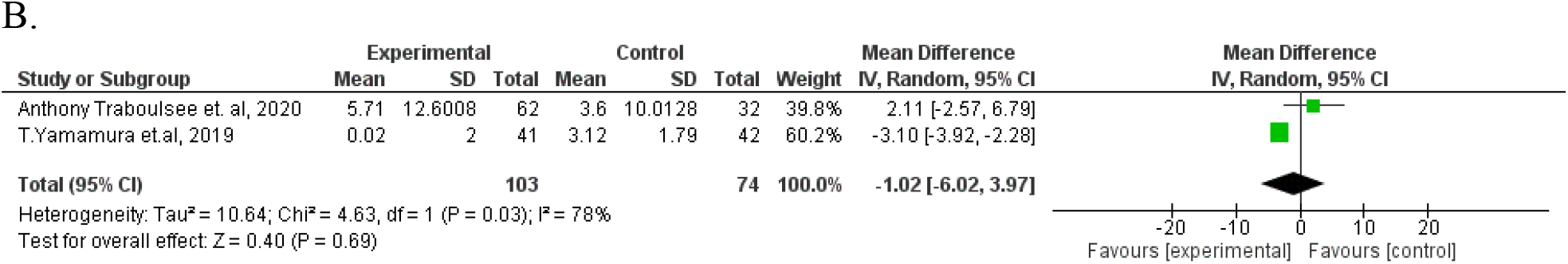
A. Change in VAS pain score B. Change in FACIT F score

#### 1. Adverse events

All included studies have reported the frequency of total adverse events as well as of serious adverse events. There was no significant difference in frequency of total reported adverse events between monoclonal antibody and the comparator arm (RR: 1.01 [0.95, 1.07], P=0.74, I^2^=14%).Similarly, the frequency of serious adverse events in both groups was not statistically different (RR: 0.83 [0.64, 1.08], P=0.16, I^2^=0%) (Figure 5A &B)

**Fig: 5.**
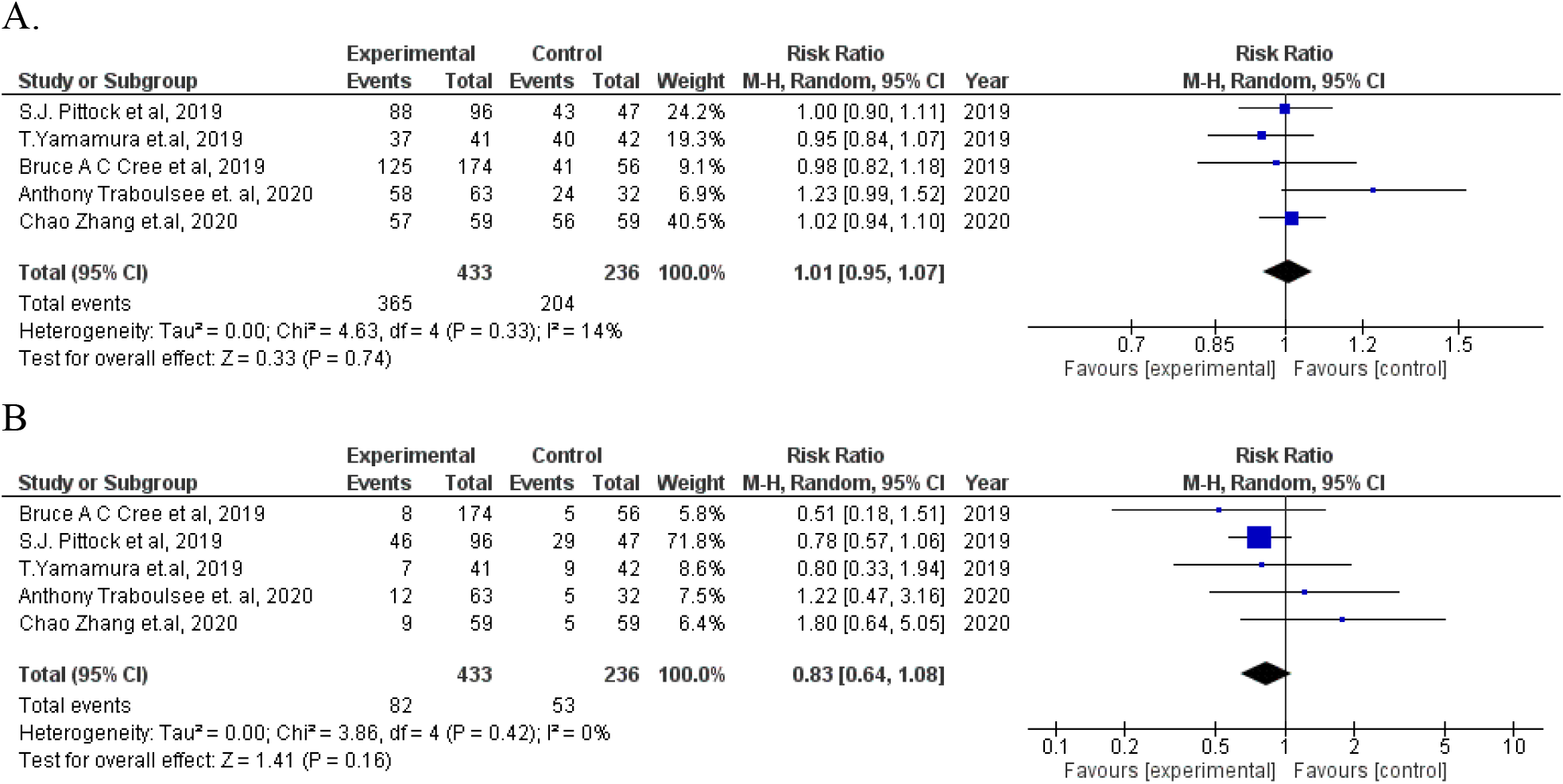
A: total adverse events B. Serious adverse events

#### 2. Other outcomes

Two studies have reported change in modified rankin scale (MrS) and EQ-5D score. No significant difference was found in both outcomes in monoclonal antibody group and control group. The results are shown in Figure 6.

**Figure: 6.**
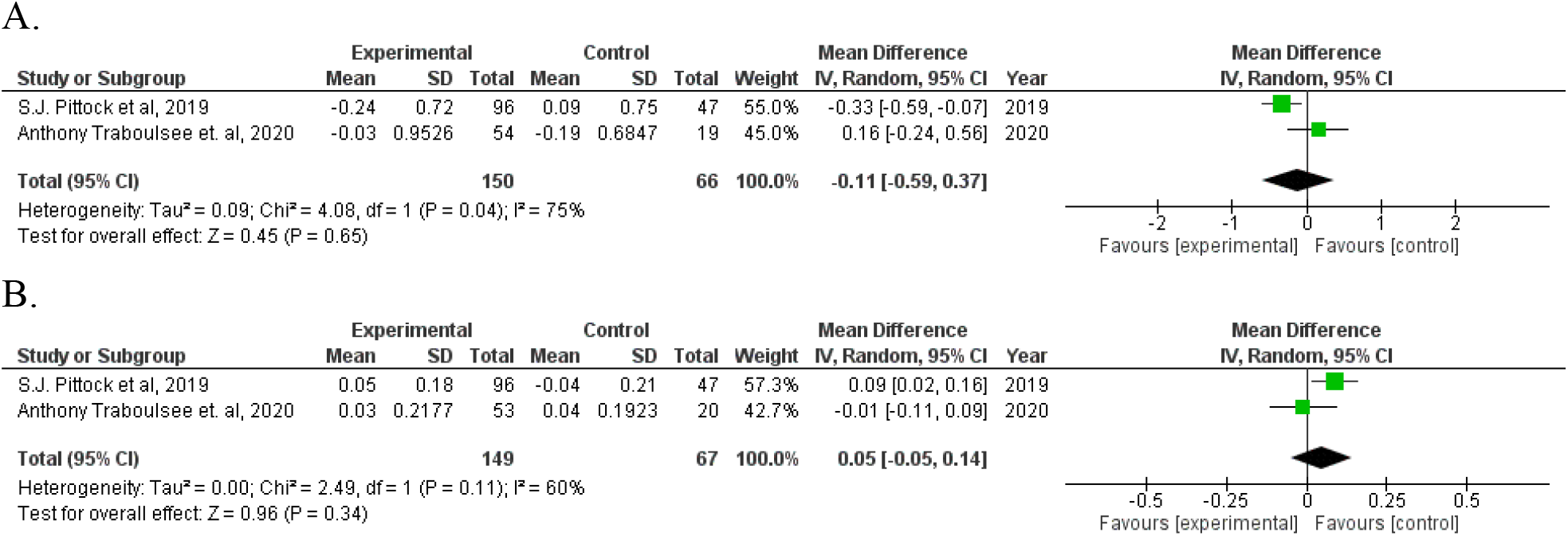
A. change in modified rankin scale B. Change in EQ 5D score

#### 3. Bias of included studies

Summary of bias from ROB 2.0 tool was generated from Revman 5.4 (Figure 7) None of the included studies had high risk of bias in any domain.

**Fig 7:**
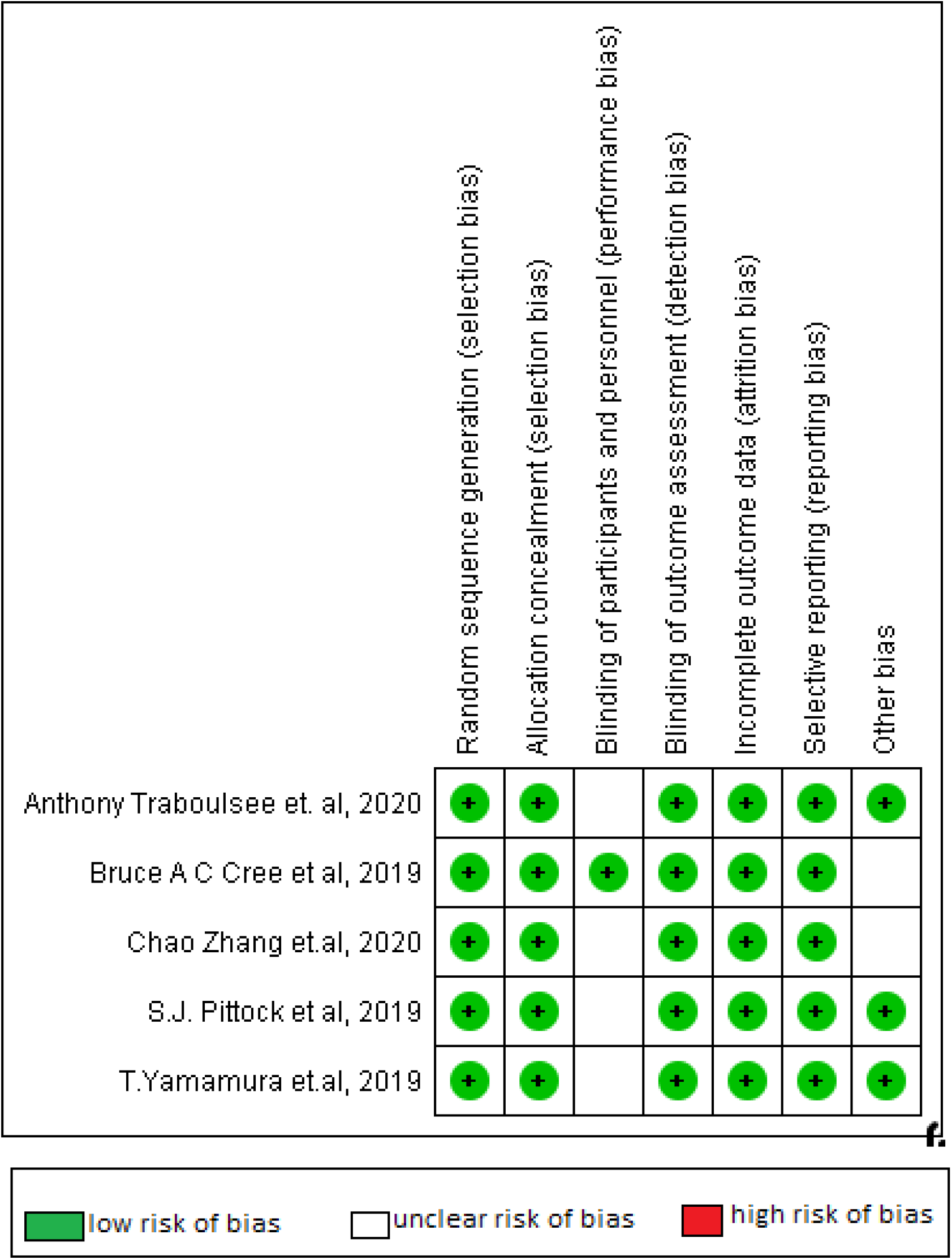
Risk of bias summary

## Discussion

It was found that all four drugs had significant benefits in relapse prevention reflected by reduced ARR and a lower frequency of relapses under treatment. The build-up of neurologic impairment caused by frequent relapses causes disability in NMOSD patients ^26^. More than half of NMOSD patients become functionally blind or wheelchair dependent if they do not receive adequate preventative therapy^27,28^. As a result, the primary goal of NMOSD therapy is to avoid illness recurrence.

This meta-analysis included low-biased RCTs and found sufficient evidence to support the use of monoclonal antibody treatment in patients with NMOSD to reduce relapse rates. The reduced relapse represents the better patients’ functional recovery. This is the probable reason why monoclonal antibody treatment can lead to a mean decrease in the EDSS score. The change in EDSS score was not altered by intervention. Tocilizumab was associated with reduction in ARR and risk of relapse that was consistent across retrospective and prospective studies. The certainty of our findings related to overall high mortality was assessed.

For AQP-4 seropositive individuals, monoclonal antibody therapy may be more effective where monoclonal antibody therapy should be routine treatment to prevent relapse. Eculizumab can be the best choice among all monoclonal antibodies at present. Aquaporumab is a newly developed recombinant human monoclonal antibody that blocks the interaction between AQP-4 and the pathogenic AQP-4 antibody. The effects of this monoclonal antibody have been tested in vitro and in vivo ^29^. This therapeutic approach may be the potential first choice in the future clinical studies.

In this systematic review and meta-analysis, efficacy and safety of monoclonal antibodies in NMO was summarised and evaluated. Despite significant variability in tocilizumab trials, exploring outcomes such as adaptive recurrence rate, change in EDSS score, and change in pain and fatigue severity scale, retrospective studies have shown that tocilizumab had positive benefits when compared to controls. Prospective studies followed a similar direction of association, though CIs were not conclusive.

The pathogenic mechanism of NMOSD, as illustrated in in vivo and in vitro researches has proved therapies targeting antibody-producing B cells are effective. Rituximab (anti-CD20) and inebilizumab (anti-CD19) which mainly act by depleting B cells had similar efficacy with other monoclonal antibodies in relapse prevention ^30^. Some other new monoclonal antibodies like ublituximab, a chimeric monoclonal antibody targeting CD20, have been proved to have a higher binding affinity to FcγRIIIa and elicit stronger antibody dependent cellular cytotoxicity compared with Rituximab ^31^. The previous clinical trials of ublituximab were mainly focused on the treatment of patients with MS (multiple sclerosis) or leukemia. An open-label, single-center, phase 1 study is already in progress to evaluate the efficacy and safety of ublituximab in NMOSD (Clinical trial.gov code: NCT02276963). Bevacizumab, another recombinant humanized monoclonal antibody binds to vascular endothelial growth factor (VEGF), plays a vital role in NMOSD^32^. An open-label, single-center, phase 1 study trial is underway to assess the efficacy and safety of bevacizumab among patients with NMOSD (Clinical trial.gov code: NCT01777412). More randomized controlled trials of ublituximab, bevacizumab, aquaporumab and other new monoclonal antibodies should be done to assess the efficacy of monoclonal antibody therapy.

This meta-analysis compared safety outcomes between monoclonal antibody group and placebo group but no significant differences in the rate of total adverse events and serious adverse effects were observed, confirming that monoclonal antibody therapy has a high level of safety in NMOSD patients. Moreover, it was found that monoclonal antibody therapy did not significantly reduce the serious adverse events. The frequencies and types of total adverse events are similar in the monoclonal antibody and placebo groups that include headache, nasopharyngitis, infection (upper respiratory tract or urinary tract), infusion-related reaction, and pain (limb, joint or back). These adverse events were caused by drug use or were accidental. In addition, Eculizumab reported to increase the risk of meningococcal sepsis in some studies^33^. In an open-label trial by Pittock et al 2013^34^, 1 in 14 participants had meningococcal sepsis after injection along with the use of Eculizumab but none had meningococcal sepsis after meningococcal vaccination in study by Pittock et al, 2019 ^35^. Rituximab and Inebilizumab may have a possible concern of elevated risk of cancer and infection^36,37^. However, according to our included RCTs, no malignancies occurred and the infection rate in monoclonal antibody group was lower than that of placebo group^21,38^. Long term follow-up in larger populations is necessary to prove the full safety profile. For Satralizumab and Tocilizumab, the main safety concern is cardiovascular disease because of the increasing blood cholesterol levels^39^. But, the incidence of cardiovascular events has not increased in monoclonal antibody group as pointed by RCT of satralizumab^40^. Although there are some potential risk factors as mentioned above, the monoclonal antibody therapy is safe based on our meta-analysis.

Multiple limitations and considerable sources of inter-study heterogeneity were highlighted in this review. The majority of included studies were non-randomized cohorts of relatively modest size. Although, most studies necessitated corticosteroids, azathioprine, rituximab, and mycophenolate, participant criteria were not entirely consistent across the studies. The dosage and delivery of monoclonal antibodies varied across non-randomized studies, and in nearly all studies, patients were on concomitant medications such as rituximab and azathioprine at the discretion of the treating physician. The causal association of specific monoclonal antibodies with outcomes was precluded.

Because the outcomes of the research were diverse, with a combination of clinical characteristics and laboratory data, a single consistent endpoint could not be reported. Also, there was no consistency in the duration of follow-up and timing of outcomes among different studies. Statistical heterogeneity that was measured by I^2^ was significant, and therefore the findings of our meta-analysis should be interpreted with caution. All the residual heterogeneity using the factors that we assessed was not explained. The outcomes were assessed against a background of concurrent use of steroids, other medications, and route of drug administration, all of which distorted the results. Both retrospective studies and RCTs were included to increase value and timeline of our review with four specific monoclonal antibodies, two primary endpoints (number of relapse and change in EDSS score) and a number of secondary endpoints. Publication bias was not significant while reporting the effects in this study. In absence of sufficient data for meta-analysis, summary outcomes were presented with qualitative synthesis to ensure the review was comprehensive. The data presented in this study represent findings from different countries and offers ethnic diversity.

## Conclusion

This meta-analysis included low-biased RCTs and found enough evidence to justify the use of monoclonal antibody therapy to minimize recurrence rates in patients with NMOSD. In contrast, the serious adverse events were not reduced with the use of monoclonal antibodies. The concomitant use of multiple medications might have altered the results. Our findings, particularly seen from the context of a few RCTs, support the pursuit of larger, multi-center RCTs that evaluate the effectiveness of each of the currently available monoclonal antibodies and better describe their adverse risk profile.

## Supporting information

Characteristics of included study

Outcomes of the included studies

Quality assessment of included studies

## Data Availability

All the data are available in the manuscript itself

## Acknowledgement

None

## Data availability statement

All the data are available in the manuscript itself

## Author’s Contribution

SS conceptualized, wrote the original manuscript, reviewed, and edited the manuscript. SS and RC were involved in study selection. SS and RC extracted the data. SG, AKP, and KD were involved in the synthesis of results. RD, BK, BSP, SD and MB were involved in the discussion of resolving disagreement in selection of the articles. SS, RC, SG, AKP, KD, BK, RD, BSP, SD and MB reviewed and edited the original manuscript.

---

**Supplementary Table 1** Quality assessment of included studies

Table 1: Characteristics of included study

Table 2: Outcomes of the included studies

