## Supplementary material for "Systematic Review and Meta-analysis of Eculizumab, Inebilizumab, Tocilizumab, and Satralizumab for Neuromyelitis Optica": Characteristics of included study

| Author and year of study | Study design | Study country | Monoclonal antibody | AQP-4 serostatus | History of relapse | Previous rituximab | Dosage regimen | Sample size | Follow up duration | Efficacy |
| --- | --- | --- | --- | --- | --- | --- | --- | --- | --- | --- |
| Manabu Araki et.al, 2014 | Observational | Japan | tocilizumab | Positive | Minimum 2 relapses in 24 months | No | 8 mg/kg | 7 | 1 year | 1. EDSS score 2. No of relapses 3. Pain and fatigue severity scale |
| Anthony Traboulsee et. l, 2020 | RCT, Phase III | Multinational | Satralizumab | Both positive and negative | At least 1 relapse in 12 months | No | 120 mg | 168 | 1.5 year | 1. EDSS score 2. No of relapses 3. Pain and fatigue severity scale |
| Chao Zhang et.al, 2020 | Open label, randomized trial, Phase II | China, multicenter | Tocilizumab | Both positive and negative | Minimum 2 relapses in 24 months | No | 8 mg/kg every 4 weeks | 118 | 60 weeks | 1. EDSS score 2. No of relapses |
| Ilya Ayzenberg et.al, 2013 | Retrospective case series | Germany | Tocilizumab | Positive | Minimum 2 relapses in 24 months | Yes | 6 mg/kg every 6 weeks in two patient and in 4 weeks in 1 patient | 3 | 1.5 year | 1. EDSS score 2. No of relapses |
| Marius Ringelstein et.al, 2015 | Retrospective observational | Germany | Tocilizumab | 6 positive out of eight | Present | Yes | 6-8 mg/kg | 8 | 10-51 months | 1. EDSS score 2. No of relapses 3. Pain severity scale |
| Itay Lotan et.al, 2019 | Retrospective | USA | Tocilizumab | Both positive and negative | Was not used as inclusion criteria | Yes (11/12) | NA | 12 | 2014-2019 | 1. No of relapses 2. Ambulatory status |
| T.Yamamura et.al, 2019 | RCT, phase III | Japan | Satralizumab | Both positive and negative | Two relapses in 2 years | No | 120 mg | 83 | 107.4 week | 1. EDSS score 2. No of relapses 3. Pain and fatigue severity scale |
| Sean J Pittock et.al, 2013 | Open label pilot study | USA | Eculizumab | Positive | Two relapses in 6 months or  Three relapse in 12 months | Yes (4/14) | 600 mg IV weekly for 4 weeks, 900 mg in 5^th^ week, and then 900 mg every 2 weeks for 48 weeks | 14 | 1 year | 1. EDSS score 2. No of relapses |
| Dean M. Wingerchuk et al, 2021 | RCT | Multinational | Eculizumab | Positive | 2 relapse in 12 months or 3 relapse in 24 months | No | 900 mg weekly for 1^st^ 4 f dose, 1200 mg every 2 weeks starting at week 4 | 143 | 211 week | 1. EDSS score 2. No of relapses |
| Bruce A C Cree et al, 2019 | RCT | Multinational | Inebilizumab | Both positive and negative | 1 relapse in 12 months or 2 relapse in 24 months | Yes | 300 mg iv at day 1 and 15 | 230 | 12 months | 1. EDSS score 2. No of relapses |
| S.J. Pittock et al, 2019 | RCT | Multinational | Eculizumab | Positive | 2 relapses in 12 months, or 3 relapses in 24 months, at least 1 occurred within the previous 12 months | No | 900 mg weekly for the first four doses starting on day 1, followed by 1200 mg every 2 weeks starting at week 4 | 143 |  | 1. EDSS score 2. No of relapses |

Table 1: Characteristics of included study
