## Supplementary material for "Systematic Review and Meta-analysis of Eculizumab, Inebilizumab, Tocilizumab, and Satralizumab for Neuromyelitis Optica": Outcomes of the included studies

| SN No | **Author and year of study** | **No of relapse** | **Change in EDSS score** | **Pain severity score** | **Fatigue severity score** | **Adverse events** |
| --- | --- | --- | --- | --- | --- | --- |
| 1 | Manabu Araki et.al, 2014 | Decreased from 2.9 ± 1.1 to 0.4 ± 0.8 (p , 0.005) | Decreased from 5.1 ± 1.7 (range, 3.0–6.5) to 4.1 ± 1.6 (range, 2.0–6.0) | Reduced from 3.0 ± 1.5 to 0.9 ± 1.2 at 12 months | Reduced from 6.1 ± 2.0 to 3.0 ± 1.4 at 12 months | upper respiratory infections, acute enterocolitis, acute pyelonephritis, leukocytopenia and/or lymphocytopenia |
| 2 | Anthony Traboulsee et. l, 2020 | Satralizumab:  19/63(30%0  Placebo: 16/32(50pct)  HR 0·45 (0·23 to 0·89); p=0·018 | Satralizumab:  53 [–0·34 (–0·62 to –0·05)]  Placebo:  20 [–0·17 (–0·52 to 0·19)] | Satralizumab:  62 [–2·74 (–11·20 to 5·73)]  Placebo:  32[ –5·95 (–15·55 to 3·65)] Difference:  3·21 (–5·09 to 11·52); p=0·44 | Satralizumab:  62 [5·71 (2·51 to 8·91)]  Placebo:  32 [3·60 (–0·01 to 7·22)]  Difference:  2·11 (–1·01 to 5·22) | urinary tract infection and upper respiratory tract infection |
| 3 | Chao Zhang et.al, 2020(RCT) | HR 0·274 [95% CI 0·123–0·607]; p=0·0006 | Relative risk 3·667 [95% CI 1·603–8·387]; p=0·0005 | NA | NA | urinary tract infection, upper respiratory tract infection, anemia, leukopenia |
| 4 | Ilya Ayzenberg et.al, 2013 | decreased from 3.0 (range, 2.3-3.0) to 0.6 (range, 0-1.3) | EDSS did not change in 2 patient and 1 patient changed EDSS from 5 to 4 | NA | NA | NA |
| 5 | Marius Ringelstein et.al, 2015 | Decreased from 4.0 (interquartile range, 3.0-5.0) to 0.4 (interquartile range, 0.0-0.8) (P = .008) | Decreased from 7.3 (interquartile range, 5.4-8.4) to 5.5 (interquartile range, 2.6-6.5) (P = .03). | Decreased from 6.5 (5.0-7.0) to 2.5 (0.3-4.5) (P = .02) | NA | nausea, gastritis, transient diarrhea, headache, fatigue, and recurrent urinary tract infections |
| 6 | Itay Lotan et.al, 2019 | Decreased from 2.0 (interquartile range = 5.75-1.29 to 0 ( interquartile range= 1.0-0) p=0.0015 | Ambulatory status:  improved in two patients remained stable in nine | NA | NA | urinary tract infections, skin abscess, asymptomatic neutropenia |
| 7 | T.Yamamura et.al, 2019 | Satralizumab:  STZ: 8/41  Placebo:  18/42  Hazard ratio:  0.34 (0.15 to 0.77) | Satralizumab:  −0.10  Placebo:  −0.21 | Satralizumab:  0.35±4.52 Placebo:  −3.73±4.12 4.08  Hazard ratio: (−8.44 to 16.61)  P=0.52 | Satralizumab:  0.02±2.00 Placebo:  3.12±1.79 Hazard ratio: −3.10 (−8.38 to 2.18) | Infection, neoplasm |
| 8 | Sean J Pittock et.al, 2013 | 3 to 0 (range zero to one) (p<0·0001) | 3·5 to 4·0 (0– 10·0) (mean diff erence 0·3 [95% CI –0·1 to 0·6]; p=0·50) | NA | NA | Headache, nausea, dizziness, diarrhea, coughing, abdominal pain |
| 9 | Dean M. Wingerchuk et al, 2021 | Eculizumab:  0.025 (95% CI = 0.013–0.048) Placebo:  0.350 (95% CI = 0.199–0.616) | Trend toward improvement no data available | NA | NA | Headache, upper respiratory tract infection, nasopharyngitis, urinary tract infection, arthralgia, back pain, diarrhea, and nausea |
| 10 | Bruce A C Cree et al, 2019 | AQP4-IgG seropositive:  Inebilizumab: 18/161 (11%) placebo 22/52(42%) (HR 0·227 [95% CI 0·121–0·423]; p<0·0001)  Intention-to-treat population:  Inebilizumab: 21/174 (12%) placebo 22/56(39%) (hazard ratio [HR] 0·272 [95% CI 0·150–0·496]; p<0·0001 | AQP4-IgG seropositive:  Placebo: 18/52 (35%) Inebilizumab:25/161(16%) OR 0·371 (0·181 to 0·763) p=0·0070  Intention-to-treat population:  Placebo: 19/56(34%) Inebilizumab: 27/174 (16%) OR 0·370 (0·185 to 0·739) 0·0049 | NA | NA | Urinary tract infection, arthralgia, back pain, headache, nasopharyngitis, diarrhoea, upper respiratory tract infection |
| 11 | S.J. Pittock et al, 2019 | Eculizumab:  3/96(3%)  Placebo:  20/47(43%)  Hazard ratio:  0.04 (0.01 to 0.15) p<0.001 | Eculizumab:  –0.18±0.81 Placebo:  0.12±0.95 –Hazard ratio:  0.29 (–0.59 to 0.01) | NA | NA | Upper respiratory tract infection, headache, nasopharyngitis, nausea, diarrhea, urinary tract infection |

Table 2: Outcomes of the included studies
