## Supplementary material for "Systematic Review and Meta-analysis of Eculizumab, Inebilizumab, Tocilizumab, and Satralizumab for Neuromyelitis Optica": Quality assessment of included studies

**Supplementary Table 1** Quality assessment of included studies.

| No | First author | Study Objectives | Study Period | Patient Selection Criteria | International  Or multi centers | Tocilizumab Treatment | baseline equivalence groups | primary outcome | follow-up period | HR with 95% CI | limitations considered | Overall score |
| --- | --- | --- | --- | --- | --- | --- | --- | --- | --- | --- | --- | --- |
| 1 | Manabu Araki et.al, 2014 | Yes | Yes | Yes | No | Yes | No | Yes | Yes | No | Yes | 7 |
| 2 | Anthony Traboulsee et. l, 2020 | Yes | Yes | Yes | Yes | Yes | Yes | Yes | Yes | Yes | Yes | 10 |
| 3 | Chao Zhang et.al, 2020 | Yes | Yes | Yes | Yes | Yes | Yes | Yes | Yes | Yes | Yes | 10 |
| 4 | Ilya Ayzenberg et.al, 2013 | Yes | Yes | Yes | No | Yes | Yes | Yes | Yes | No | Yes | 8 |
| 5 | Ankelien Duchow et.al, 2021 | Yes | Yes | Yes | No | Yes | Yes | Yes | Yes | No | Yes | 8 |
| 6 | Marius Ringelstein et.al, 2015 | Yes | Yes | Yes | No | Yes | Yes | Yes | Yes | No | Yes | 8 |
| 7 | Itay Lotan et.al, 2019 | Yes | Yes | Yes | No | Yes | Yes | Yes | Yes | No | Yes | 8 |
| 8 | T.Yamamura et.al, 2019 | Yes | Yes | Yes | No | Yes | Yes | Yes | Yes | Yes | Yes | 9 |
| 9 | Sean J Pittock et.al, 2013 | Yes | Yes | Yes | No | Yes | Yes | Yes | Yes | No | Yes | 8 |
| 10 | Dean M. Wingerchuk | Yes | Yes | Yes | Yes | Yes | Yes | Yes | Yes | No | Yes | 9 |
| 11 | Bruce A C Cree et al, 2019 | Yes | Yes | Yes | Yes | Yes | Yes | Yes | Yes | Yes | Yes | 10 |
| 12 | S.J. Pittock et al, 2019 | Yes | Yes | Yes | Yes | Yes | Yes | yes | No | Yes | Yes | 9 |
